# Reward Processing in Children with Psychotic-like Experiences

**DOI:** 10.1101/2021.06.01.21257966

**Authors:** Jasmine Harju-Seppänen, Haritz Irizar, Elvira Bramon, Sarah-Jayne Blakemore, Liam Mason, Vaughan Bell

## Abstract

Alterations to striatal reward pathways have been identified in individuals with psychosis. They are hypothesised to be a key mechanism that generate psychotic symptoms through the production of aberrant attribution of motivational salience and are proposed to result from accumulated childhood adversity and genetic risk, making the striatal system hyper-responsive to stress. However, few studies have examined whether children with psychotic-like experiences (PLEs) also exhibit these alterations, limiting our understanding of how differences in reward processing relate to hallucinations and delusional ideation in childhood. Consequently, we examined whether PLEs and PLE-related distress were associated with reward-related activation in the nucleus accumbens (NAcc). The sample consisted of children (N = 6,718) from the Adolescent Brain Cognitive Development (ABCD) study aged 9-10 years who had participated in the Monetary Incentive Delay (MID) task in functional MRI. We used robust mixed-effects linear regression models to investigate the relationship between PLEs and NAcc activation during the reward anticipation and reward outcome stages of the MID task. Analyses were adjusted for gender, household income, ethnicity, depressive symptoms, movement in the scanner, pubertal development, scanner ID, subject and family ID. There was no reliable association between PLEs and alterations to anticipation-related or outcome-related striatal reward processing. We discuss the implications for developmental models of psychosis and suggest a developmental delay model of how PLEs may arise at this stage of development.

## Introduction

Psychotic-like experiences (PLEs) include delusion-like beliefs and hallucinations that remain below the threshold for psychotic disorder. PLEs are relatively common in childhood (Linscott & Os, 2013) with a median prevalence of 17% among children aged 9–12 years and 7.5% among those aged 13–18 years (Kelleher et al., 2012). Even during childhood however, they are a predictor of later transition to psychosis (Fisher et al., 2013; Healy et al., 2019; Poulton et al., 2000) and poor physical and mental health outcomes across the lifespan (Davies et al., 2018; Healy et al., 2019; Kelleher et al., 2012; Trotta et al., 2020).

It is unclear how childhood PLEs relate to established neurocognitive mechanisms for adult psychosis. One hypothesis is that both adult and childhood PLEs are associated with altered striatal dopamine (MacManus et al., 2012). In the developmental risk factor model, dysregulated striatal dopamine is cited as a final common pathway where genetic risk and developmental adversity converge and lead to psychosis via the generation of aberrant salience (Howes & Kapur, 2009; Murray et al., 2017) – a process where typically innocuous experiences are assigned heighted motivational salience due to misfiring of striatal dopamine leading to delusions and perceptual aberrations. Indeed, neuroimaging studies on reward processing in psychosis risk states and prodromal periods indicate that dysregulation of striatal dopamine is detectable before the onset of frank psychotic disorder (reviewed in Howes et al., 2020). Initial evidence suggests that there is an earlier association between dysregulated striatal reward processing and psychotic-like experiences in 14-19 year-old adolescents (Papanastasiou et al., 2018).

However, it is still not clear if altered striatal reward processing that predicts psychosis and psychosis-risk in older individuals would necessarily explain the presentation of PLEs at a younger age. This is important because childhood is a crucial point of risk divergence for PLEs. An estimated 75–90% of psychotic experiences during childhood and adolescence are transitory (Rubio et al., 2012; van Os et al., 2009) but those whose PLEs do not resolve have particularly poor outcomes (Calkins et al., 2017; Downs et al., 2013) with distress related to PLEs at age 12 adding predictive value for poor outcome later in life (Sullivan et al., 2020). However, PLEs at ages 8-15 years show a weaker relationship with later poor outcome than PLEs at ages 16 and over, despite a greater prevalence at this earlier age (Schimmelmann et al., 2015), suggesting they may not fully reflect the same mechanism as PLEs in later adolescence. Consequently, understanding whether dysregulated reward processing is associated with PLEs and PLE-related-distress during earlier childhood could provide important evidence to understand to what extent these experiences reflect an early disruption to a key causal mechanism present in later psychosis. The aim of the current study was therefore to examine whether PLEs and PLE-related distress in childhood is associated with alterations to striatal reward processing.

Reward processing consists of both reward anticipation and reward evaluation and that these functions of the reward system dissociate (Schultz, 2002). The monetary incentive delay (MID) task was designed to distinguish these functions when used in functional magnetic imaging studies (fMRI) studies (Knutson et al., 2000). It has been used extensively in psychosis research and meta-analysis of relevant fMRI studies provide strong evidence for striatal reward system dysregulation in adults with frank psychosis (Radua et al., 2015). Additional studies have also found evidence for these alterations in antipsychotic naïve patients with schizophrenia(Nielsen et al., 2012), and in adults with PLEs (Wotruba et al., 2014). Evidence from concurrent fMRI and positron emission tomography (PET) indicates that changes to dopamine transport underlie changes in fMRI reward system-related activation during the MID (Dubol et al., 2018), suggesting that fMRI studies of the MID task are a reliable proxy for alterations to reward-related dopamine function. Studies with children indicate that the paradigm is valid for measuring reward processing in this age group (Dougherty et al., 2018).

Small sample sizes and lack of representative sampling are a challenge for fMRI studies(Turner et al., 2018) and this has been cited as a particular issue for neuroimaging studies of children (Herting et al., 2018). Here, we aimed to test whether PLEs or PLE-related distress was associated with dysregulated reward-processing in the left and right nucleus accumbens (NAcc) during childhood by examining the association between activation during the fMRI MID task in a large (N=6,900+) sample of 9–10-year-olds, who were part of the Adolescent Brain Cognitive Development (ABCD) study (Garavan et al., 2018). The ABCD study is an ongoing cohort study including more than 11,000 children and includes extensive social, cognitive and developmental measures. It includes demographics, measures of PLEs and PLE-related distress and, in over half of children, the MID task in fMRI.

Consequently, we aimed to test whether PLEs or PLE-related distress could be explained by dysregulated striatal reward processing by examining the association between NAcc activation during the fMRI monetary incentive delay task in a large (N=6,500+) sample of 9– 10-year-olds. We tested both anticipation-related and outcome-related reward processing in the left and right NAcc while controlling for potential confounders.

## Methods

### Sample

The ABCD dataset (release 3.0; https://abcdstudy.org/) includes 11,878 children aged 9-10 years (Volkow et al., 2018). This is a longitudinal dataset being collected at 21 sites across the US. Full details of recruitment are described in Garavan et al. (2018). Institutional review board approval was obtained for each site before data collection and all parents provided written informed consent in addition to assent from the participants (Clark et al., 2018).

Data from participants was excluded based on the following criteria: having a psychiatric diagnosis (N = 1973), not completing the Prodromal Questionnaire (N = 12), taking psychotropic medication (N = 1032), not completing the MID task in the scanner (N = 1030), insufficient performance on the task (N= 573), missing motion data (N = 455), missing fMRI data (N = 27), and if reward-related activation in the NAcc was more than three standard deviations from the mean (N = 223 for reward anticipation; N = 123 for reward outcome), leaving N = 6,718 who contributed to either the final anticipation or outcome analysis.

### Measures

### Psychotic-like experiences

Psychotic-like experiences (PLEs) were measured using the Prodromal Questionnaire – Brief Child Version, a modified version of the Prodromal Questionnaire Brief Version (PQ-B)(Loewy et al., 2011) – a self-report measure for psychosis risk syndromes that has been validated in nine to ten year olds (Karcher et al., 2018). Unlike the Kiddie Schedule for Affective Disorders and Schizophrenia (KSADS-5), also available in the ABCD dataset, the PQ-BC allows for measurement of PLEs alongside a measure of distress for the same items. The PQ-BC is a 21-item questionnaire that measures unusual perceptions and sensations, ideas of reference, affective changes, unusual beliefs, or abnormally suspicious thoughts, along with associated distress. The PQ-BC consists of two parts: the first asks whether the individual has had any of the listed psychotic-like thoughts, feelings and experiences, with an overall score ranging from 0-21. If they answer yes, participants also indicate how related distressing in the second part (from 1-5). A subset of six items were selected to represent analogues of positive symptoms of psychosis (thought interference, visual hallucination, auditory hallucination, two items for paranoia and bizarre beliefs). PLE types were derived from this variable, where participants were categorised as having no PLEs, non-distressing PLEs, or distressing PLEs. Additional analyses used total sum of PLEs and PLE-related distress.

### Depressive symptomology

Depressive symptoms were measured using the Kiddie Schedule for Affective Disorders and Schizophrenia (KSADS-5). The ABCD study used a recently validated and computerised version of the KSADS-5 (Townsend et al., 2020). The following depressive symptoms were added to create a depression score: depressed mood, anhedonia, and irritability.

### Pubertal development

Pubertal development was measured using the Pubertal Development Scale (PDS) (Petersen et al., 1988). Child-provided data was used as the primary measure of pubertal stage and missing scores were supplemented by parent-reported information.

### Monetary Incentive Delay task

The Monetary Incentive Delay (MID) task (Knutson et al., 2000) measures the anticipation and receipt of rewards and losses. Participants are presented with an incentive cue (2000ms) at the beginning of each trial (Win $5, Win $0.20, Lose $0.20, Lose $5 or $0-no money at stake), followed by a jittered anticipation event (lasting 1500-4000ms). Participants then need to respond to a variable target (150-500ms), in order to either win or avoid losing money. In the ABCD study, participants are presented with 40 reward (20 small reward, 20 large reward) and 40 loss anticipation trials (20 small loss, 20 large loss), 20 no money anticipation trials, and feedback trials(Casey et al., 2018). The task was individualised with the initial duration of the response target drawn from a practice session completed by the participant prior to entering the scanner. In order to reach a 60% accuracy rate, task difficulty was adjusted during the task after every third incentivised trial based on the overall accuracy rate of the previous six trials. The target duration was shortened if the individual’s accuracy fell below the target accuracy level. Participants who did not reach acceptable performance in the task were excluded from analysis (indexed by whether all trial types resulted in more than three events for both positive and negative feedback), as well as those whose NAcc activity was above or below three standard deviations. The MID task has been previously validated in typically developing children during fMRI (Cao et al., 2019) and validation studies for the paradigm and data used in this study have been previously described in Casey et al. (2018) and Chaarani et al. (2021). Casey et al. (2018) reported that the experimental manipulation was successful in maintaining hit rates at close to 60%, and that reaction times and payoff amounts were consistent across experimental runs. Chaarani et al. (2021) reported that the task is associated with robust brain activations which are consistent with the extant literature.

### Imaging acquisition

The primary outcome was reward-related activation during the MID task from the left and right NAcc. Full details on imaging acquisition is reported in Casey et al. (2018). Imaging data was collected across sites using multi-channel coils and multiband echo planar imaging acquisition. Scanning included a fixed order of localiser, T1- and T2-weighted images, resting state, and diffusion-weighted imaging. Three tasks (MID task, stop signal, and emotional n-back) were completed in an order randomised across participants. Blood□oxygen□level□dependent (BOLD) images were acquired using gradient EPI with standardised acquisition parameters.

### Imaging processing and analysis

The ABCD Data Analysis and Informatics Center performed centralised processing and analysis of the imaging data. Full information regarding this is detailed in Hagler et al.(Hagler et al., 2019), and is summarised here. Left and right NAcc regions of interest were derived from subcortical segmentation using FreeSurfer 5.3.0 (Fischl, 2012). Estimated task-related activation were computed for individual subjects using the general linear model in AFNI 3dDeconvolve and were available as contrast beta weights. The contrasts used in this study were “large reward vs. no money” and “small reward vs. no money” for reward anticipation activity, and “all reward positive vs. negative feedback” for reward outcome activity. For these contrasts, region of interest average beta coefficients were computed for each of the two runs and then averaged.

### Statistical analysis

We conducted analyses to investigate the association between PLEs, presence of any non-distressing PLEs, and presence of any distressing PLEs, and reward-related activation in the NAcc using multi-level regression analyses across the population sample. Each analysis was conducted and reported separately for two outcomes: left and right NAcc activation during the reward anticipation stage of the MID task, and left and right NAcc activation during reward outcome stage of the MID task. We tested for evidence of heteroscedasticity in the data, and due to its presence, estimated the effects of the predictor variables using robust mixed effects linear regression models (Koller, 2016).

For all analyses, we initially tested for a minimally adjusted association between PLE type and reward-related activation, adjusted only for the random effects covariates (subject ID, nested within family ID, and scanner ID). We then subsequently updated the model to include additional fixed effect covariates to test the association after adjustment for potential confounders. These included sex, household income, parental education, ethnicity, motion in scanner, depressive symptoms and pubertal development.

Sex, household income, parental education, and ethnicity were included as potential confounders owing to their association with psychosis risk (Jongsma et al., 2020; Morgan et al., 2009; Murphy et al., 2020). Depression was included as a potential confounder due to its association with alterations in reward processing (Keren et al., 2018). Pubertal development was included as a potential confounder due to associations between reward processing and puberty (Ladouceur et al., 2019). Motion in scanner was included due known role as a confounder in fMRI activation studies. Missing data for the covariates was imputed through multiple imputation using the *Mice* package in R (Buuren & Groothuis-Oudshoorn, 2011). Polytomous regression was used for unordered factor variables. Proportional odds model was used for ordered factor variables. Logistic regression imputation was used for binary variables.

We subsequently repeated the main analyses but included all individuals with psychiatric diagnoses and medication known to have significant impact on reward processing (stimulants and anti-psychotics, see supplementary tables 1-4). We also completed alternative analysis where PLEs were included as sum total, along with their associated distress (see supplementary tables 5-8). Finally, we performed analyses only in individuals with a psychiatric diagnosis (see supplementary tables 9-10).

All analyses were conducted in *R* (version 3.6.2) using the *robustlmm* package (Koller, 2016). The data was transformed into long format using *reshape* (Wickham, 2007) to allow us to test for multivariate outcomes. All analysis code and analysis output for this study has been made freely available on an Open Science Framework archive: https://osf.io/vqzhu/?view_only=3851605a5ab74267ab68b35c207ef90a

## Results

After applying our exclusion criteria, 6718 participants remained for either the reward anticipation (N = 6553) and/or reward outcome (N = 6654) analyses. The demographic characteristics of the sample are shown in Table 1.

**Table 1.** Demographic characteristics of participants who contributed to either the reward anticipation or outcome analysis (N= 6718)

|  | *Total*<br>(N = 6718) | *No PLEs*<br>(N = 4025) | *Non-distressing*<br>*PLEs*<br>(N = 833) | *Distressing*<br>*PLEs*<br>(N = 1860) |
| --- | --- | --- | --- | --- |
| *Age (SD)* | 9.9 (0.62) | 9.9 (0.62) | 9.9 (0.62) | 9.9 (0.62) |
| *Gender N (%)* |  |  |  |  |
| Female | 3494 (52.0%) | 2107 (52.3%) | 386 (46.3%) | 1001 (53.8%) |
| *Household income (USD)* |  |  |  |  |
| <50k | 1872 (27.9%) | 984 (24.4%) | 236 (28.3%) | 652 (35.1%) |
| >=50k & <100k | 1866 (27.8%) | 1090 (27.1%) | 237 (28.4%) | 539 (29.0%) |
| >=100k | 2980 (44.4%) | 1951 (48.5%) | 360 (43.2%) | 669 (36.0%) |
| *Parental education N (%)* |  |  |  |  |
| < HS diploma | 398 (5.9%) | 181 (4.5%) | 49 (5.9%) | 168 (9.0%) |
| HS diploma/ GED | 641 (9.5%) | 347 (8.6%) | 76 (9.1%) | 218 (11.7%) |
| Some college | 1847 (27.5%) | 989 (24.6%) | 262 (31.5%) | 596 (32.0%) |
| Bachelor | 2002 (29.8%) | 1276 (31.7%) | 230 (27.6%) | 496 (26.7%) |
| Postgraduate degree | 1830 (27.2%) | 1232 (30.6%) | 216 (25.9%) | 382 (20.5%) |
| *Ethnicity* |  |  |  |  |
| Asian | 172 (2.6%) | 114 (2.8%) | 22 (2.6%) | 36 (1.9%) |
| Black | 886 (13.2%) | 437 (10.9%) | 124 (14.9%) | 325 (17.5%) |
| Other/mixed | 1193 (17.8%) | 647 (16.1%) | 166 (19.9%) | 380 (20.4%) |
| White | 4467 (66.5%) | 2827 (70.2%) | 521 (62.5%) | 1119 (60.2%) |

### Effect of PLEs and distress on reward anticipation

As can be seen from Tables 2 and 3, there were main effects of reward magnitude and laterality on reward anticipation activity, indicating the validity of the paradigm, even after adjustment for potential confounders. However, there was no association with non-distressing or distressing PLEs in either analysis. Effects of PLE type on NAcc activation for reward anticipation are displayed by left and right laterality in Figure 1.

**Table 2.** Minimally adjusted regression model (N = 6553) examining the effect of presence and type of PLEs, reward magnitude and laterality on nucleus accumbens (NAcc) response to reward anticipation

| *Predictor* | *Estimate* | *95% CIs* | *p value* |
| --- | --- | --- | --- |
| Non-distressing PLEs | -0.001 | -0.015, 0.013 | 0.879 |
| Distressing PLEs | -0.006 | -0.017, 0.004 | 0.244 |
| Laterality (Right NAcc > Left NAcc) | -0.012 | -0.017, -0.008 | <0.001 |
| Reward magnitude (Large Reward > Small Reward) | 0.072 | 0.068, 0.077 | <0.001 |

**Table 3.** Fully adjusted regression model (N = 6553) examining the effect of presence and type of PLEs, reward magnitude and laterality on nucleus accumbens (NAcc) response to reward anticipation

| *Predictor* | *Estimate* | *95% CIs* | *p value* |
| --- | --- | --- | --- |
| Non-distressing PLEs | 0.0005 | -0.014, 0.015 | 0.949 |
| Distressing PLEs | -0.002 | -0.012, 0.009 | 0.755 |
| Laterality (Right NAcc > Left NAcc) | -0.012 | -0.017, -0.008 | <0.001 |
| Reward Magnitude (Large Reward > Small Reward) | 0.072 | 0.068, 0.077 | <0.001 |
| Gender | 0.001 | -0.009, 0.010 | 0.912 |
| Depressive symptoms | -0.005 | -0.019, 0.009 | 0.461 |
| Household income [<50K] | -0.001 | -0.015, 0.013 | 0.856 |
| Household income [≥50K & <100K] | -0.001 | -0.013, 0.010 | 0.798 |
| Parental education - < HS Diploma | -0.023 | -0.046, -0.0001 | 0.049 |
| Parental education - HS Diploma/GED | -0.025 | -0.044, -0.007 | 0.007 |
| Parental education - Post Graduate Degree | -0.007 | -0.019, 0.005 | 0.231 |
| Parental education - Some College | 0.002 | -0.015, 0.011 | 0.755 |
| Race - Asian | -0.020 | -0.049, 0.008 | 0.162 |
| Race - Black | -0.003 | -0.018, 0.012 | 0.698 |
| Race - Other | -0.005 | -0.017, 0.008 | 0.464 |
| Motion | -0.036 | -0.057, -0.016 | <0.001 |
| Pubertal development | -0.005 | -0.015, 0.005 | 0.338 |

**Figure 1.**
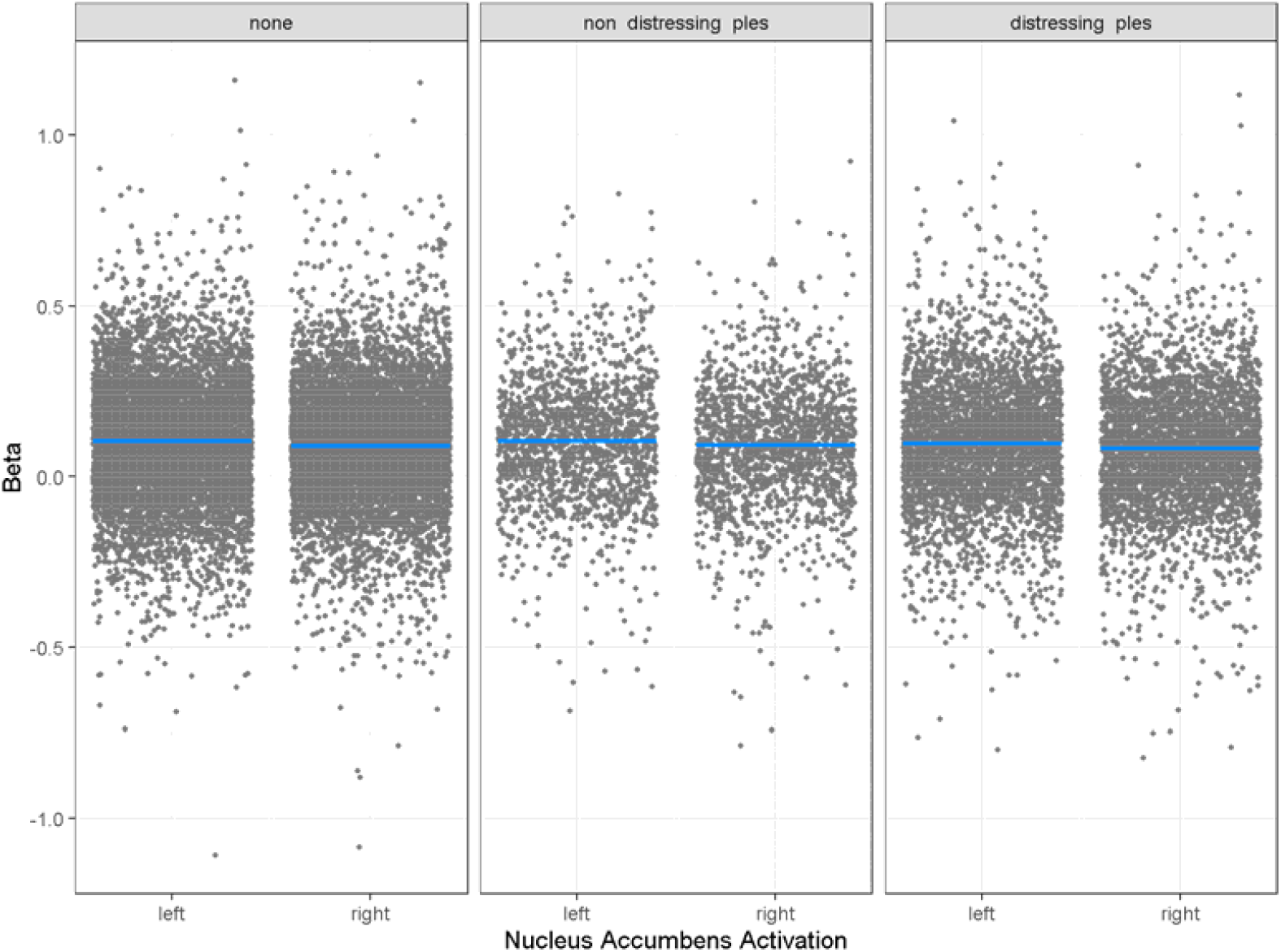
Relationship between PLE group status on left and right nucleus accumbens activation in the reward-anticipation component of the Monetary Incentive Delay task

### 2) Effect of PLEs and distress on reward outcome

As show in Tables 4 and 5, there were main effects of NAcc laterality on reward anticipation activity, even after adjustment for potential confounders, but no association with non-distressing or distressing PLEs in either analysis. Effects of PLE type on NAcc activation for reward outcome are displayed by left and right laterality in Figure 2.

**Table 4.** Minimally adjusted regression model (N = 6654) on association between types of PLEs, distress, laterality on nucleus accumbens (NAcc) response to reward outcome.

| *Predictor* | *Estimate* | *95% CIs* | *p value* |
| --- | --- | --- | --- |
| Non distressing PLEs | -0.001 | -0.015, 0.013 | 0.879 |
| Distressing PLEs | -0.006 | -0.017, 0.004 | 0.244 |
| Laterality (Right NAcc > Left NAcc) | -0.025 | -0.030, -0.021 | <0.001 |

**Table 5.** Fully adjusted regression model (N = 6654) on association between types of PLEs, distress, laterality on nucleus accumbens (NAcc) response to reward outcome.

| *Predictor* | *Estimate* | *95% CIs* | *p value* |
| --- | --- | --- | --- |
| Non-distressing PLEs | -0.005 | -0.022, 0.012 | 0.553 |
| Distressing PLEs | -0.008 | -0.021, 0.005 | 0.213 |
| Laterality (Right NAcc > Left NAcc) | -0.025 | -0.030, -0.021 | <0.001 |
| Gender | 0.008 | -0.003, 0.019 | 0.132 |
| Depressive symptoms | 0.009 | -0.007, 0.026 | 0.265 |
| Household income [<50K] | 0.002 | -0.015, 0.019 | 0.812 |
| Household income [>=50K & <100K] | -0.00002 | -0.013, 0.014 | 0.981 |
| Parental education - < HS Diploma | -0.009 | -0.037, 0.018 | 0.512 |
| Parental education - HS Diploma/GED | -0.006 | -0.029, 0.016 | 0.569 |
| Parental education - Post Graduate Degree | -0.006 | -0.021, 0.008 | 0.415 |
| Parental education - Some College | -0.002 | -0.017, 0.014 | 0.831 |
| Race - Asian | 0.022 | -0.013, 0.058 | 0.212 |
| Race - Black | -0.025 | -0.044, -0.007 | 0.007 |
| Race - Other | -0.012 | -0.027, 0.003 | 0.127 |
| Motion | 0.075 | 0.052, 0.098 | <0.001 |
| Pubertal development | 0.002 | -0.011, 0.014 | 0.806 |

**Figure 2.**
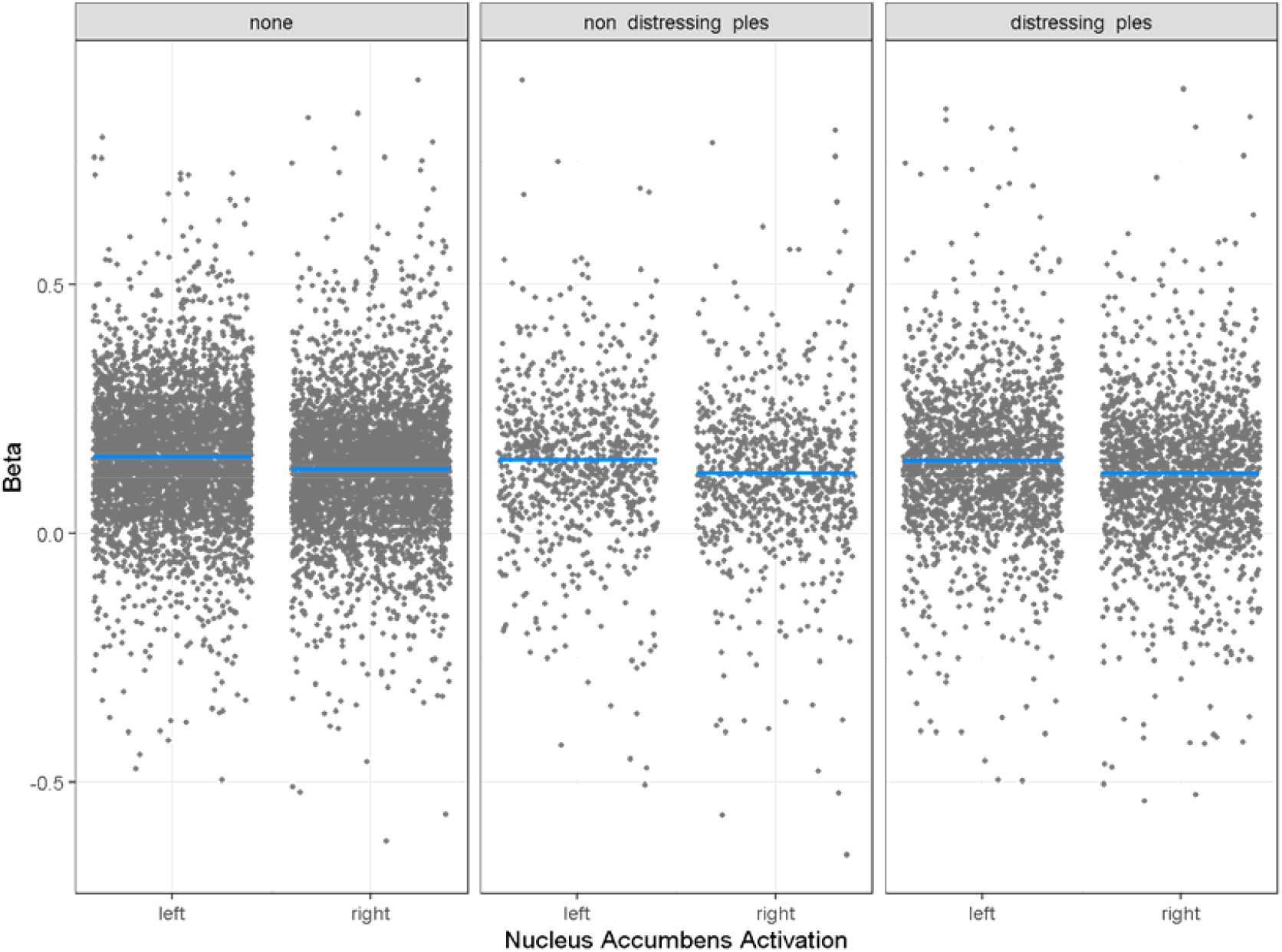
Relationship between PLE group status on left and right nucleus accumbens activation in the reward-outcome component of the MID task

In addition, we completed sensitivity analyses reported in supplementary tables 1 to 4 that included all individuals, including those with a psychiatric diagnosis or medication use. The pattern of results was similar across analyses with regard to PLEs. The only exception was that PLE-related distress was significantly associated with striatal activation in the minimally adjusted reward anticipation analysis (β = -0.013, 95% CI = -0.022, -0.004, *p* = 0.005).However, this relationship became non-significant in the fully adjusted analysis (β = -0.013, 95% CI = 0.018, 0.0001, *p* = 0.053).

We also completed additional analysis examining the effect of PLEs and PLE-related distress by coding them as sum total variables: total number of PLEs and total levels of PLE-related distress. Total number of PLEs was not related to anticipation-related reward activation (β = <0.001, 95% CI = -0.006, 0.007, *p* = 0.892) or reward outcome-related reward activation (β = 0.001, 95% CI = -0.007, 0.009, *p* = 0.855) in minimally adjusted analyses. Similarly, total level of PLE-related distress was not related to anticipation-related reward activation (β = -0.002, 95% CI = -0.005, 0.0002], *p* = 0.072) or reward outcome-related reward activation (β = -0.001, 95% CI = -0.004, 0.002, *p* = 0.460) in minimally adjusted analyses. This pattern of relationships remained unchanged in the fully adjusted analyses (full details are given in supplementary tables 5-8). We also performed additional analyses including only the subset of individuals with a psychiatric diagnosis (supplementary tables 9-10). These analyses indicated that there was no reliable association between PLEs and reward outcome. In this same subset of participants, distressing PLEs (but not non-distressing PLEs) were associated with reward anticipation activation only, showing an association with a small reduction in reward-anticipation-related activity (β = -0.033, 95% CI = -0.055, -0.012, *p* = 0.003).

## Discussion

In this large study of over 6,500 9-10-year-old children, we report no association between PLEs and NAcc activation during either the reward anticipation or reward outcome stages of an fMRI MID task. This was the case regardless of whether PLEs were included as a categorical or continuous variable. We completed a number of further supplementary analyses, including all individuals who were previously excluded due to the exclusion criteria, including PLE-related distress as a continuous measure, and including only participants with a psychiatric diagnosis. In these further analyses, only one statistical association was found: distressing PLEs (but not non-distressing PLEs) were associated with NAcc activation during reward anticipation (but not reward outcome) in the sub-population of participants with psychiatric diagnoses. Given that this result was the only significant association from a large number of tests, was a small effect, was conducted as an exploratory analysis, and was not present for PLEs without distress in the same analysis, we suggest it is unlikely to be strong evidence for the presence of this mechanism.

The findings have several implications for developmental models of psychosis risk and our understanding of explanatory mechanisms for psychosis-spectrum experiences more broadly. In terms of the psychosis risk, the developmental risk factor model (Murray et al., 2017) suggests that accumulated childhood and social adversity combined with genetic risk makes the striatal dopamine system hyper-responsive to stress. According to the model, these alterations generate the symptoms of psychosis through a process of aberrant assignment of salience to stimuli that would normally appear to have low levels of motivational significance (Howes & Kapur, 2009; Kapur, 2003). These findings have been supported by fMRI studies reporting dysregulated reward processing in the MID task in adults with psychosis (Juckel et al., 2006; Kirschner et al., 2018) and in adolescents with PLEs (Papanastasiou et al., 2018).

This study found no evidence for the presence of this mechanism at a peak age for PLEs in 9-10-year-old children despite a very large sample and validated measures that have produced reliable evidence for this association in older age groups. We also found no strong evidence for this when we included children with psychiatric diagnoses, who typically share a greater number of risk factors for reward system sensitisation. This is despite the fact that PLEs in children of this age predict poor outcome over the lifespan (Calkins et al., 2017; Downs et al., 2013), poorer cognitive abilities (Reichenberg et al., 2010) and greater levels of adversity (Trotta et al., 2015). This suggests that PLEs at this age are markers of adverse development and / or psychopathology but are potentially not associated with alterations to striatal activation, raising doubts over whether they share a mechanism proposed for psychotic-spectrum experiences later in life.

Notably, pre-adolescents have been shown to have several perceptual and reasoning differences that alter and stabilise during adolescence, potentially suggesting other mechanisms that might generate PLEs. For example, there is evidence that pre-adolescent children may perform auditory functions more unreliably than adults, tending to rely more heavily on top-down interpretation of sounds (Moore, 2012). Similarly, in the visual domain, pre-adolescent children tend to rely more on high spatial frequencies to extract local facial features to perceive fearful facial expressions whereas adolescent children use rapid decoding of global features using in the low spatial frequency ranges (Peters & Kemner, 2017). Additionally and relevant to the measurement of unusual beliefs, magical thinking is common in childhood although declines into adolescence and this is largely understood in terms of the under-development of causal reasoning (Muentener & Bonawitz, 2018) involving the understanding of transfer of physical force between objects, the outcomes of goal-directed actions produced by dispositional agents, and the ability to track covariation relations between events. Development in each of these domains may additionally be affected by developmental adversities, potentially giving rise to PLEs during preadolescence that are generated by distinct mechanisms fom PLEs reported later in life.

Based on current findings, we hypothesise that PLEs in preadolescence may be generated by delayed development of perceptual and causal reasoning. Speculatively, the presence of childhood adversity might impact the typical developmental trajectory of perceptual and causal reasoning systems, meaning that greater numbers of PLEs largely reflect developmental delay in these systems rather than dysregulation of striatal dopamine at this age. Nevertheless, accumulation of pre-existing and environmental risk factors, particularly those that have a broader impact on development (Dominguez et al., 2010), will make later hyper-responsiveness of the striatal dopamine system more likely. This interactive developmental model might account for the contrasting trajectories of PLEs from childhood to adolescence where the majority of children show resolving or attenuated PLEs and only a small high-risk minority show persistent or intermittent PLEs (Thapar et al., 2012).

However, this study presents several limitations that warrant caution when interpreting its results. One limitation is the extent to which blood-oxygen-level-dependent (BOLD) signal in the ventral striatum allows accurate localisation of dysregulated subcortical dopamine. Meta-analytic evidence from PET studies suggests that it may be the dorsal rather than ventral striatum where dopamine dysregulation may be most apparent in adult psychosis (McCutcheon et al., 2018). The area of interest used in this study was the NAcc, based in the ventral striatum. Nevertheless, activation in this area during the MID task is reliably associated with psychosis across meta-analysis of multiple studies (Radua et al., 2015). Therefore, it is likely that BOLD signal response reliably reflects altered dopamine-mediated reward processing, but it may not accurately localise it. Indeed, a prior multimodal PET-fMRI study of the MID task (Dubol et al., 2018) reported that reward anticipation was reliably associated with BOLD signal in the NAcc with PET imaging showed it was associated with dopamine transporter availability in the midbrain. Consequently, we presume it is unlikely that there would be marked dysregulation to functionally adjacent areas in the dopamine system that would not result in BOLD detectable NAcc activation during the MID task, meaning it likely remains a useful measure of altered dopamine-mediated reward processing. However, given these validation studies were conducted in adults, we also note the limitations of generalising this assumption to 9-10-year-old children. Given that PET studies are typically used to cross-validate the role a dopamine in regional fMRI activation and are not routinely conducted on children except for clinical reasons, this may be difficult to directly test, but remains a possibility.

Although the ABCD endeavoured to obtain a nationally representative sample, children from higher income families were over-represented. We attempted to address this by including family and site as random effects as suggested by Heeringa & Berglund (2020), although it is possible that this did not fully eliminate sampling biases. PLEs were measured with the Prodromal Questionnaire – Brief Child Version. This is a self-report questionnaire and although has been well validated in this sample (Karcher et al., 2018) may not have had the same sensitivity as structured interview assessments.

We note the sole positive statistical association reported in this study was between reward anticipation and distressing PLEs in children grouped by having any psychiatric diagnosis. We also note that this finding was small and seemingly very selective – it was not present for reward outcome and was not associated with non-distressing PLEs in the same analysis. However, it is possible that this group represents a subgroup where the earliest effects of dopamine system dysregulation may be found, potentially related more broadly to psychopathology, and this may be worth noting as a hypothesis for future investigation.

In conclusion, we found no reliable evidence that the presence of PLEs in a large and well-powered sample predicted dysregulated reward processing. As the ABCD study is ongoing cohort study, future research should focus on exploring the timing of PLE-related differences in reward processing arise during development.

## Data Availability

Data used in the preparation of this article were obtained from the Adolescent Brain Cognitive DevelopmentSM (ABCD) Study (https://abcdstudy.org), held in the NIMH Data Archive (NDA). This is a multisite, longitudinal study designed to recruit more than 10,000 children age 9-10 and follow them over 10 years into early adulthood. The ABCD Study is supported by the National Institutes of Health and additional federal partners under award numbers U01DA041022, U01DA041028, U01DA041048, U01DA041089, U01DA041106, U01DA041117, U01DA041120, U01DA041134, U01DA041148, U01DA041156, U01DA041174, U24DA041123, U24DA041147, U01DA041093, and U01DA041025. A full list of supporters is available at https://abcdstudy.org/federal-partners.html. A listing of participating sites and a complete listing of the study investigators can be found at https://abcdstudy.org/Consortium_Members.pdf. ABCD consortium investigators designed and implemented the study and/or provided data but did not necessarily participate in analysis or writing of this report. This manuscript reflects the views of the authors and may not reflect the opinions or views of the NIH or ABCD consortium investigators.
The ABCD data repository grows and changes over time. The ABCD data used in this report came from doi: 10.15154/1521353. DOIs can be found at https://doi.org

## Acknowledgements

Data used in the preparation of this article were obtained from the Adolescent Brain Cognitive Development^SM^ (ABCD) Study (https://abcdstudy.org), held in the NIMH Data Archive (NDA). This is a multisite, longitudinal study designed to recruit more than 10,000 children age 9-10 and follow them over 10 years into early adulthood. The ABCD Study® is supported by the National Institutes of Health and additional federal partners under award numbers U01DA041022, U01DA041028, U01DA041048, U01DA041089, U01DA041106, U01DA041117, U01DA041120, U01DA041134, U01DA041148, U01DA041156, U01DA041174, U24DA041123, U24DA041147, U01DA041093, and U01DA041025. A full list of supporters is available at https://abcdstudy.org/federal-partners.html. A listing of participating sites and a complete listing of the study investigators can be found at https://abcdstudy.org/Consortium_Members.pdf. ABCD consortium investigators designed and implemented the study and/or provided data but did not necessarily participate in analysis or writing of this report. This manuscript reflects the views of the authors and may not reflect the opinions or views of the NIH or ABCD consortium investigators.

The ABCD data repository grows and changes over time. The ABCD data used in this report came from doi: 10.15154/1521353. DOIs can be found at https://doi.org

## Funding

JHS: MRC doctoral studentship. HI: EU Horizon 2020 under a Marie Sklodowska-Curie grant (747429); National Institute of Allergy and Infectious Diseases. EB: NIHR UK (NIHR200756); Mental Health Research UK; ESRC; BMA Margaret Temple Fellowship; MRC New Investigator and Centenary Awards (G0901310 and G1100583). NIHR Biomedical Research Centre at UCL Hospitals and UCL. SJB: Wellcome (grant number WT107496/Z/15/Z); Jacobs Foundation; Wellspring Foundation; University of Cambridge. LM: MRC Clinician Scientist Fellowship (MR/S006613/1). VB is funded by the UKRI Global Challenges Research Fund.

## Notes

### Competing Interest Statement

The authors have declared no competing interest.

### Funding Statement

JHS is supported by a Medical Research Council doctoral studentship. HI has received funding from the European Union's Horizon 2020 research and innovation programme under the Marie Skoldowska-Curie grant agreement no. 747429 and is currently supported by a grant from the National Institute of Allergy and Infectious Diseases (National Institutes of Health). EB has received the following support: National Institute of Health Research UK (NIHR200756). Mental Health Research UK John Grace QC Scholarship 2018. ESRC co-funded award. BMA Margaret Temple Fellowship 2016. Medical Research Council New Investigator and Centenary Awards (G0901310 and G1100583). MRC (G1100583). NIHR Biomedical Research Centre at University College London Hospitals NHS Foundation Trust and University College London. SJB is funded by: Wellcome (grant number WT107496/Z/15/Z). The Jacobs Foundation. The Wellspring Foundation and the University of Cambridge. LM was supported by an MRC Clinician Scientist Fellowship (MR/S006613/1). VB is funded by the UKRI Global Challenges Research Fund.

### Author Declarations

Ethical approval for data collection and dissemination of the data for secondary data analysis was approved by the Institutional Review Board (cIRB) at the University of California, San Diego and is fully described in Auchter et al. 2018 (doi: 10.1016/j.dcn.2018.04.003). In line with this approval, access to the data and its use in this study was granted to the authors following an application to the National Institute of Health (NIH). The study registration is available at the following URL: https://eur01.safelinks.protection.outlook.com/?url=https%3A%2F%2Fdoi.org%2F10.15154%2F1521353&data=04%7C01%7C%7C14a7dbd712a14ed48a0b08d924c0cf77%7C1faf88fea9984c5b93c9210a11d9a5c2%7C0%7C0%7C637581232802409882%7CUnknown%7CTWFpbGZsb3d8eyJWIjoiMC4wLjAwMDAiLCJQIjoiV2luMzIiLCJBTiI6Ik1haWwiLCJXVCI6Mn0%3D%7C1000&sdata=1F1YukfYpGNLtzLf0C82S1rbMVvOwvnaqUDpiyKqMaE%3D&reserved=0

