## Supplementary material for "Reward Processing in Children with Psychotic-like Experiences"

**Supplementary Table 1:** Sensitivity analysis – Minimally adjusted regression model including all individuals (N = 8808) examining the effect of presence and type of PLEs, reward magnitude and laterality on nucleus accumbens (NAcc) response to reward anticipation.

| *Predictor* | *Estimate* | *95% CIs* | *p value* |
| --- | --- | --- | --- |
| Non-distressing PLEs | -0.002 | -0.014, 0.011 | 0.808 |
| Distressing PLEs | -0.013 | -0.022, -0.004 | 0.0055 |
| Laterality (Right NAcc > Left NAcc) | -0.011 | -0.015, -0.008 | <0.001 |
| Reward Magnitude (Small Reward > Large Reward) | -0.070 | -0.074, -0.066 | <0.001 |

**Supplementary Table 2:** Sensitivity analysis – Fully adjusted regression model including all individuals (N = 8808) examining the effect of presence and type of PLEs, reward magnitude and laterality on nucleus accumbens (NAcc) response to reward anticipation.

| *Predictor* | *Estimate* | *95% CIs* | *p value* |
| --- | --- | --- | --- |
| Non-distressing PLEs | -0.001 | -0.013, 0.011 | 0.840 |
| Distressing PLEs | -0.010 | -0.0184, 0.0001 | 0.053 |
| Laterality (Right NAcc > Left NAcc) | -0.012 | -0.0155, -0.0080 | <0.001 |
| Reward magnitude (Small Reward > Large Reward) | -0.070 | -0.0737, -0.0663 | <0.001 |
| Gender | 0.002 | -0.0057, 0.0100 | 0.589 |
| Depressive symptoms | -0.003 | -0.0144, 0.0087 | 0.631 |
| Household income [<50K] | 0.003 | -0.0092, 0.0150 | 0.639 |
| Household income [>=50K & <100K] | 0.005 | -0.0047, 0.0151 | 0.305 |
| Parental education - < HS Diploma | -0.020 | -0.0396, -0.0004 | 0.046 |
| Parental education - HS Diploma/GED | -0.023 | -0.0389, -0.0072 | 0.004 |
| Parental education - Post Graduate Degree | -0.006 | -0.0169, 0.0042 | 0.237 |
| Parental education - Some College | -0.005 | -0.0163, 0.0057 | 0.343 |
| Race - Asian | -0.005 | -0.0313, 0.0217 | 0.723 |
| Race - Black | 0.0001 | -0.0128, 0.0130 | 0.989 |
| Race - Other/Mixed | -0.004 | -0.0150, 0.0065 | 0.440 |
| Motion | -0.047 | -0.0638, -0.0307 | <0.001 |
| Pubertal development | -0.009 | -0.0177, 0.0001 | 0.052 |
| Diagnosis - schizophrenia | -0.139 | -0.3491, 0.0720 | 0.197 |

**Supplementary Table 3:** Sensitivity analysis – Minimally adjusted regression model including all individuals (N = 8939) on association between types of PLEs, distress and laterality on nucleus accumbens (NAcc) response to reward outcome.

| *Predictor* | *Estimate* | *95% CIs* | *p value* |
| --- | --- | --- | --- |
| Non-distressing PLEs | 0.00001 | -0.015, 0.015 | 0.999 |
| Distressing PLEs | -0.004 | -0.015, 0.006 | 0.417 |
| Laterality (Right NAcc > Left NAcc) | -0.027 | -0.031, -0.023 | <0.001 |

**Supplementary Table 4:** Sensitivity analysis – Fully adjusted regression model including all individuals (N = 8939) on association between types of PLEs, distress and laterality on nucleus accumbens (NAcc) response to reward outcome.

| *Predictor* | *Estimate* | *95% CIs* | *p value* |
| --- | --- | --- | --- |
| Non-distressing PLEs | -0.002 | -0.016, 0.013 | 0.840 |
| Distressing PLEs | -0.007 | -0.018, 0.004 | 0.230 |
| Laterality (Right NAcc > Left NAcc) | -0.027 | -0.031, -0.023 | <0.001 |
| Gender | 0.007 | -0.003, 0.016 | 0.175 |
| Depressive symptoms | 0.015 | 0.001, 0.029 | 0.030 |
| Household income [<50K] | -0.001 | -0.016, 0.014 | 0.890 |
| Household income [>=50K & <100K] | -0.001 | -0.012, 0.011 | 0.928 |
| Parental education - < HS Diploma | -0.007 | -0.031, 0.016 | 0.544 |
| Parental education - HS Diploma/GED | -0.007 | -0.026, 0.012 | 0.449 |
| Parental education - Post Graduate Degree | -0.001 | -0.014, 0.012 | 0.893 |
| Parental education - Some College | -0.001 | -0.014, 0.013 | 0.928 |
| Race - Asian | 0.015 | -0.018, 0.048 | 0.368 |
| Race - Black | -0.027 | -0.042, -0.011 | <0.001 |
| Race - Other/Mixed | -0.006 | -0.019, 0.007 | 0.330 |
| Motion | 0.068 | 0.050, 0.087 | <0.001 |
| Pubertal development | -0.0002 | -0.011, 0.010 | 0.972 |
| Diagnosis - schizophrenia | -0.480 | -0.734, -0.225 | <0.001 |

**Supplementary Table 5:** Minimally adjusted regression model (N = 6553) on association between number of PLEs and total distress, reward magnitude and laterality on nucleus accumbens (NAcc) response to reward anticipation.

| *Predictor* | *Estimate* | *95% CI* | *p value* |
| --- | --- | --- | --- |
| Number of PLEs | 0.0005 | -0.006, 0.007 | 0.883 |
| Laterality (Right NAcc > Left NAcc) | -0.012 | -0.017, -0.008 | <0.0001 |
| Total distress | -0.002 | -0.005, 0.0002 | 0.068 |
| Reward magnitude (Large Reward > Small Reward) | -0.072 | -0.077, -0.068 | <0.0001 |

**Supplementary Table 6:** Fully adjusted regression model (N = 6553) on association between number of PLEs and total distress, reward magnitude and laterality on nucleus accumbens (NAcc) response to reward anticipation.

| *Predictor* | *Estimate* | *95% CI* | *p value* |
| --- | --- | --- | --- |
| Number of PLEs | 0.001 | -0.005, 0.008 | 0.718 |
| Laterality (Right NAcc > Left NAcc) | -0.012 | -0.017, -0.008 | <0.001 |
| Total distress | -0.002 | -0.005, 0.001 | 0.119 |
| Reward magnitude (Large Reward > Small Reward) | -0.072 | -0.077, -0.068 | <0.001 |
| Gender | 0.0001 | -0.009, 0.009 | 0.977 |
| Depressive symptoms | -0.003 | -0.017, 0.012 | 0.727 |
| Household income [<50K] | -0.001 | -0.015, 0.013 | 0.882 |
| Household income [>=50K & <100K] | -0.001 | -0.012, 0.010 | 0.842 |
| Parental education - < HS Diploma | -0.022 | -0.045, 0.001 | 0.056 |
| Parental education - HS Diploma/GED | -0.025 | -0.043, -0.007 | 0.008 |
| Parental education - Post Graduate Degree | -0.008 | -0.020, 0.004 | 0.212 |
| Parental education - Some College | -0.002 | -0.015, 0.011 | 0.777 |
| Race - Asian | -0.021 | -0.049, 0.008 | 0.153 |
| Race - Black | -0.002 | -0.017, 0.013 | 0.775 |
| Race - Other/Mixed | -0.004 | -0.017, 0.008 | 0.499 |
| Motion | -0.036 | -0.056, -0.016 | <0.001 |
| Pubertal development | -0.004 | -0.015, 0.006 | 0.404 |

**Supplementary Table 7:** Minimally adjusted regression model (N = 6554) on association between number of PLEs, total distress and laterality on nucleus accumbens (NAcc) response to reward outcome.

| *Predictor* | *Estimate* | *95% CI* | *p value* |
| --- | --- | --- | --- |
| Number of PLEs | 0.001 | -0.007, 0.009 | 0.855 |
| Laterality (Right NAcc > Left NAcc) | -0.025 | -0.030, -0.021 | <0.001 |
| Total distress | -0.001 | -0.004, 0.002 | 0.460 |

**Supplementary Table 8:** Fully adjusted regression model (N = 6554) on association between number of PLEs, total distress and laterality on nucleus accumbens (NAcc) response to reward outcome.

| *Predictor* | *Estimate* | *95% CI* | *p value* |
| --- | --- | --- | --- |
| Number of PLEs | -0.0005 | -0.008, 0.007 | 0.908 |
| Laterality (Right NAcc > Left NAcc) | -0.025 | -0.030, -0.021 | <0.001 |
| Total distress | -0.001 | -0.004, 0.002 | 0.546 |
| Gender | 0.008 | -0.003, 0.019 | 0.143 |
| Depressive symptoms | 0.010 | -0.007, 0.026 | 0.255 |
| Household income [<50K] | 0.002 | -0.015, 0.019 | 0.806 |
| Household income [>=50K & <100K] | 0.0002 | -0.013, 0.014 | 0.976 |
| Parental education - < HS Diploma | -0.010 | -0.037, 0.018 | 0.492 |
| Parental education - HS Diploma/GED | -0.006 | -0.029, 0.016 | 0.569 |
| Parental education - Post Graduate Degree | -0.006 | -0.020, 0.009 | 0.423 |
| Parental education - Some College | -0.002 | -0.017, 0.013 | 0.803 |
| Race - Asian | 0.022 | -0.013, 0.058 | 0.214 |
| Race - Black | -0.025 | -0.043, -0.007 | 0.008 |
| Race - Other/Mixed | -0.012 | -0.027, 0.003 | 0.127 |
| Motion | 0.075 | 0.052, 0.098 | <0.001 |
| Pubertal development | 0.002 | -0.011, 0.014 | 0.808 |

**Supplementary Table 9:** Minimally adjusted regression model including only individuals with a psychiatric diagnosis (N = 1325) examining the effect of presence and type of PLEs, reward magnitude and laterality on nucleus accumbens (NAcc) response to reward anticipation.

| *Predictor* | *Estimate* | *95% CIs* | *p value* |
| --- | --- | --- | --- |
| Non-distressing PLEs | -0.011 | -0.042, 0.020 | 0.487 |
| Distressing PLEs | -0.033 | -0.055, -0.012 | 0.003 |
| Laterality (Right NAcc > Left NAcc) | -0.010 | -0.029, -4.36E-05 | 0.049 |
| Reward Magnitude (Large Reward > Small Reward) | 0.054 | 0.044, 0.064 | <0.001 |

**Supplementary Table 10:** Minimally adjusted regression model including only individuals with a psychiatric diagnosis (N = 1311) examining the effect of presence and type of PLEs, and laterality on nucleus accumbens (NAcc) response to reward outcome.

| *Predictor* | *Estimate* | *95% CIs* | *p value* |
| --- | --- | --- | --- |
| Non-distressing PLEs | 0.023 | -0.016, 0.062 | 0.245 |
| Distressing PLEs | 0.002 | -0.020, 0.029 | 0.898 |
| Laterality (Right NAcc > Left NAcc) | -0.026 | -0.037, -0.015 | <0.001 |
